# GASTRIC ACIDITY IN DIFFERENT PHENOTYPES OF SYMPTOMATIC GASTROESOPHAGEAL REFLUX DISEASE

**DOI:** 10.1101/2023.04.11.23288425

**Authors:** Jerry D. Gardner

## Abstract

**Background:** The present report examines gastric pH from normal subjects and different gastroesophageal reflux disease (GERD) phenotypes to compare the distributions of gastric pH values and of changes in gastric acid concentrations to those reported previously for esophageal pH from the same groups of subjects. I also examined total esophageal acidity as a function of total gastric acidity in the different groups of subjects.

**Methods:** I analyzed 24-hour gastric and esophageal pH recordings from normal subjects and subjects with a particular GERD phenotype to calculate total integrated acidity and total time pH<4. I also examined gastric pH recordings for the distributions of gastric pH values and the distributions of changes in gastric acidity.

**Results:** There were different distributions for gastric pH, but virtually identical distributions of changes in gastric acid concentrations in the different groups of subjects. There was a significant positive relationship between total integrated esophageal acidity and total integrated gastric acidity in different GERD phenotypes, but not in normal subjects. The slope of the line relating integrated esophageal acidity to gastric acidity correlated directly with the responses of different GERD phenotypes to PPI treatment reported previously by others.

**Conclusions:** It seems possible esophageal acidity and gastric acidity can influence each other and by so doing account for the variation in the differences in the distributions of values of pH and acid concentrations among normal subjects and different GERD phenotypes. Furthermore, the strength of the relationship between esophageal and gastric acidity can determine the response to PPI treatment.

## INTRODUCTION

Previously (1), I reported that in analyses of time-series data for esophageal pH from normal subjects and subjects with a particular GERD phenotype, values for esophageal pH followed a power law distribution for each group of subjects. The distribution of changes in esophageal acid concentration spanned four orders of magnitude and variation was greatest in normal subjects, less in Functional Heartburn subjects, still less in Reflux Hypersensitivity subjects and least in NERD subjects.

Since gastric acid is the source of esophageal acid, I have conducted the same analyses in the same subjects in whom I previously analyzed esophageal pH (1). I have also examined the extent to which the relationship between esophageal acidity and gastric acidity might account for the reported clinical response of different GERD phenotypes to treatment with a proton pump inhibitor (PPI).

## SUBJECTS

Subjects for the present analyses were the same subjects used for previous analyses (1-4).

Patients were identified by exploring the electronic database at the Royal London Hospital GI Physiology Unit that contains clinically indicated impedance-pH recordings (Sandhill Scientific, Highlands Ranch, CO) from patients with typical symptoms of gastroesophageal reflux.

Using Lyon consensus definitions of symptomatic GERD phenotypes,(5, 6), I selected 24-hour esophageal and gastric pH recordings from normal subjects (n=20), Functional Heartburn subjects (n=20), Reflux Hypersensitivity subjects (n=20), and nonerosive esophageal reflux disease (NERD) subjects (n=20). All subjects had a normal upper gastrointestinal endoscopy at the time of the impedance-pH study. The pH recordings from one normal subject were technically unsatisfactory and were omitted from the present analyses.

As reported previously (1, 3), for this retrospective analysis of clinically indicated tests with no identifiable patient data, the Stanford University Institutional Review Board determined that this research does not involve human subjects as defined in 45 CFR 46.102(f) or 21 CFR 50.3 (g) (7).

Values from impedance-pH testing from subjects for the present analyses have been published previously (1-4). Normal subjects, Functional Heartburn subjects and Reflux Hypersensitivity subjects all had normal esophageal acid exposure time (AET) with normal esophageal pH <4 for less than 4% of the 24-hour esophageal pH recording. NERD subjects had increased esophageal AET of pH <4 for greater than 6% of the 24-hour esophageal pH recording. Reflux Hypersensitivity subjects had a positive association of symptoms with reflux episodes (Symptom Index (8)) and a Symptom-association Probability (9), whereas Functional Heartburn subjects had no association of symptoms with reflux episodes (negative Symptom Index and negative Symptom-association Probability). Normal subjects had no symptoms during the impedance-pH testing.

## METHODS

Methods for the present analyses were identical to those used for previous analyses (1-3).

The impedance-pH catheter was inserted with an esophageal pH sensor positioned 5cm above the upper border of the lower esophageal sphincter and a gastric pH sensor positioned 15cm below the esophageal pH sensor. Six impedance channels were positioned 3, 5, 7, 9, 15 and 17cm above the lower esophageal sphincter, respectively. Software provided by Sandhill to process pH recordings automatically adjusts all pH values for the difference between the calibration temperature of 25C and the recording temperature of 37C. Software provided by Sandhill was also used to export pH data for every 4^th^ second of the recording to an Excel file.

All pH values below 0.5 were replaced by 0.5 and all pH values above 7.5 were replaced by 7.5.

To determine cumulative acid concentration for a subject, all pH values were converted to acid concentration in mmol/L The sum of all values for acid concentration for that subject was calculated and each value of acid concentration was then expressed as a percentage of the cumulative acid concentration. The distribution of the change in sequential values of acid concentration was calculated for gastric acidity for each subject. The frequency distributions of changes in acid concentration for each group of subjects were calculated as the means of the values from all subjects in the group for each bin of the distribution. Expressing acid concentration as a percentage of cumulative acid concentration for a given subject makes it possible to examine differences in the distributions of changes in acid concentration that do not depend on the magnitude of the acid concentration per se. Others (10) have calculated the change in values in a time-series as a percentage difference from the mean of all values.

Total acidity was calculated as integrated acidity described previously (11) for each esophageal and gastric pH record as the time-weighted average of the acid concentration in mmol/L. Total acidity was also calculated as the percentage of the total time pH<4.

Curve fitting and statistical analyses were performed using GraphPad Prism 9.4.1 software. Because the present analyses were exploratory, P-values were not adjusted for multiple comparisons.

## RESULTS

Figure 1 displays the distributions of gastric pH values from normal subjects and GERD phenotypes. Results in Table 1 show that each distribution in Figure 1 is significantly non-linear by a Runs test and is significantly better fit by a 5^th^-order polynomial than by a 4^th^ order polynomial. Each distribution was not significantly better fit by a 6^th^-order polynomial than by a 5^th^ order polynomial by an F-test (Results not shown).

**Figure 1.**
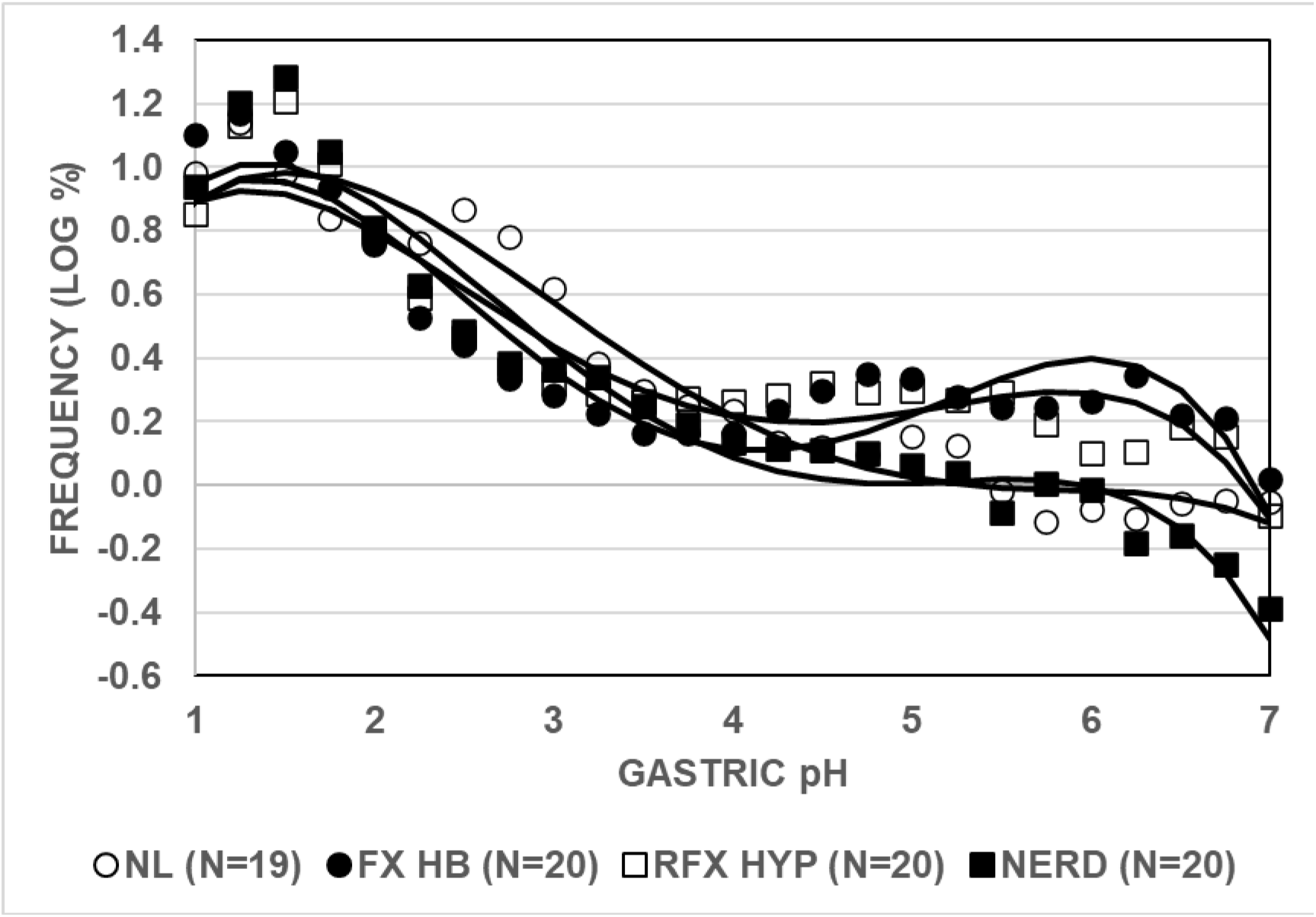
Distributions of values of gastric pH in normal subjects and GERD phenotypes. Values given on the Y-axis are mean frequencies for the bin indicated on the X-axis. The solid lines are from a fit of the data to a fifth-order polynomial. Abbreviations are NL – normal subjects; FX HB – Functional Heartburn subjects; RFX HYP – Reflux Hypersensitivity subjects; NERD – Nonerosive Reflux Disease subjects. The number of subjects in each group is given in parentheses after each abbreviation.

**Table 1.** RESULTS OF CURVE FITTING OF DISTRIBUTIONS OF GASTRIC pH VALUES IN NORMAL SUBJECTS AND GERD PHENOTYPES.

| PHENOTYPE | P-VALUE SLOPE NON-LINEAR | 5<sup>TH</sup> ORDER POLY BETTER FIT THAN 4<sup>TH</sup> ORDER POLY | F-VALUE (DFn, DFd) |
| --- | --- | --- | --- |
| NL | 0.0006 | 0.0168 | 6.75 (1, 21) |
| FX HB | 0.0001 | <0.0001 | 60.68 (1, 21) |
| RFX HYP | 0.0024 | 0.018 | 6.59 (1, 21) |
| NERD | 0.0027 | 0.0003 | 18.27 (1, 21) |

Abbreviations are NL – normal; FX HB – Functional Heartburn; RFX HYP – Reflux Hypersensitivity; NERD – non erosive reflux disease; POLY - polynomial. The p-value for non-linear slope is from a Runs test. The p-value comparing polynomial fits is from an F-test DFn – degrees of freedom numerator; DFd – degrees of freedom denominator.

The polynomial model has no physiologic significance. It is simply a model that creates a curve that comes close to the data points and makes it possible to test the data for statistical differences as summarized in Table 2.

**Table 2.** COMPARISON OF THE POLYNOMIAL FITS OF DISTRIBUTIONS OF GASTRIC pH VALUES IN NORMAL SUBJECTS AND GERD PHENOTYPES.

|  |  |  |
| --- | --- | --- |
| **COMPARISON** | **P-VALUE** | **F-VALUE (DFn, DFd)** |
| ARE SLOPES DIFFERENT? | <0.0001 | 6.58 (18, 84) |
| NL VS FX HB | <0.0001 | 14.88 (6, 42) |
| NL VS RFX HYP | 0.0026 | 4.07 (6, 42) |
| NL VS NERD | 0.0051 | 3.66 (6, 42) |
| FX HB VS RFX HYP | 0.1648 | 1.62 (6, 42) |
| FX HB VS NERD | <0.0001 | 17.16 (6, 42) |
| RFX HYP VS NERD | 0.0003 | 5.54 (6, 42) |
| Abbreviations are NL – normal; FX HB – Functional Heartburn; RFX HYP – Reflux Hypersensitivity; NERD – non erosive reflux disease; DFn – degrees of freedom numerator; DFd – degrees of freedom denominator. The p-value is from an F-test. |  |  |

Results in Table 2 show that each pair of comparisons was significantly different except the pair comparing the distribution of gastric pH values from Functional Heartburn subjects to that from Reflux Hypersensitivity subjects.

Previously, I found that the distribution of changes in cumulative esophageal acidity is biphasic for each GERD phenotype and normal subjects (1). Each distribution spanned at least 4 orders of magnitude, and was best fit by a 6^th^ order polynomial (1). Furthermore, all pairwise comparisons were significantly different at P<0.0001 by an F-test, and the variation in esophageal acid concentrations was greatest in normal subjects, less in Functional Heartburn, still less in Reflux Hypersensitivity and least in NERD (1). In contrast to the distributions of changes in esophageal acid concentrations, the distributions of changes in gastric acid concentrations illustrated in Figure 2, although biphasic, span only 1.5 orders of magnitude, and are essentially identical for all GERD phenotypes and normal subjects.

**Figure 2.**
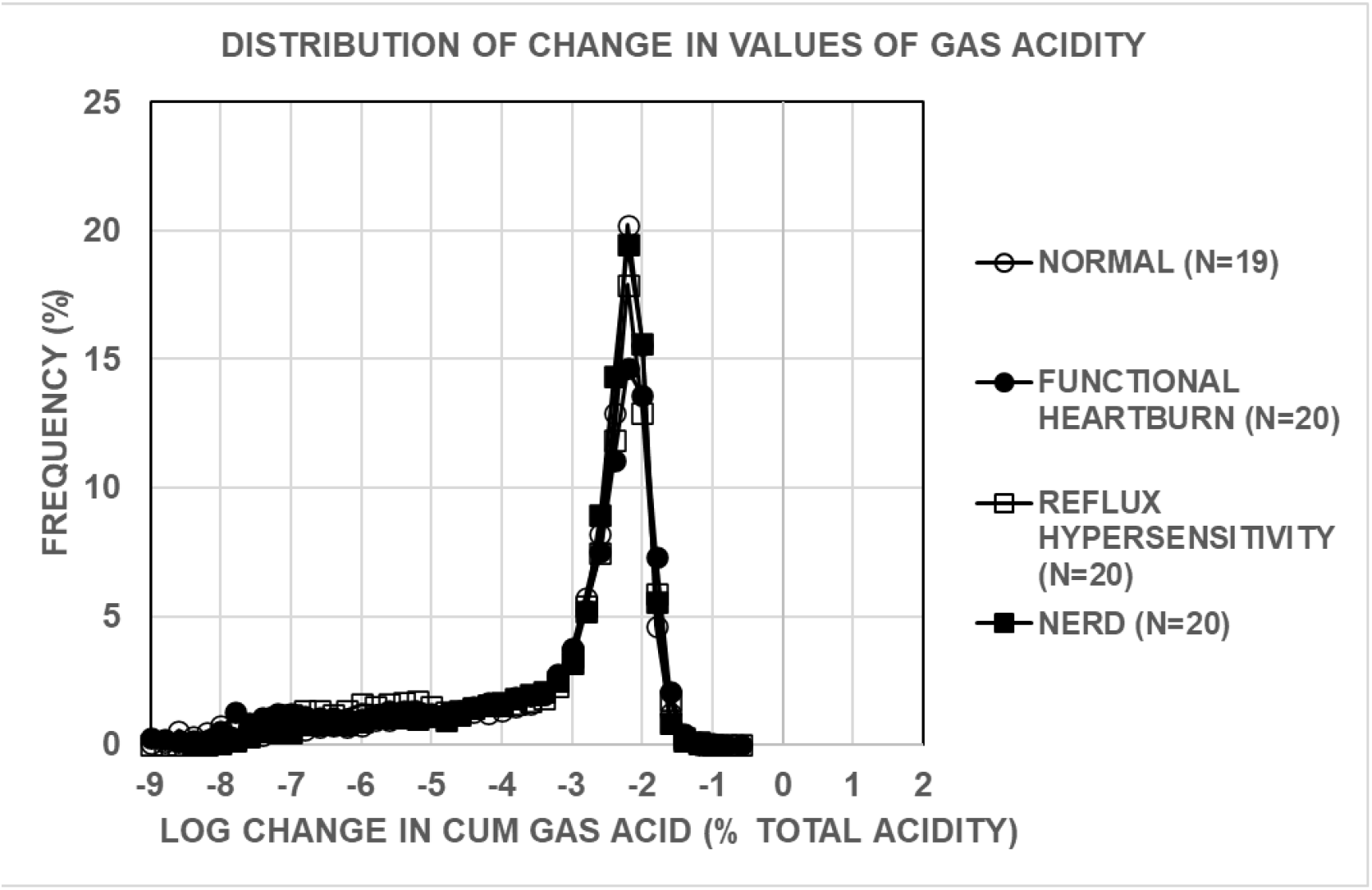
Distributions of change in values for cumulative gastric acidity for GERD phenotypes and normal subjects. Values given on the Y-axis are mean frequencies for the bin indicated on the X-axis. Abbreviations are CUM – cumulative; GAS – gastric.

Results in Table 4 show that the slope of the line relating total integrated esophageal acidity to total integrated gastric acidity in Figure 3 was significantly different from zero for Reflux Hypersensitivity and NERD subjects but nor for Functional Heartburn and Normal subjects. The slope was significantly non-linear for Normal subjects but not for any GERD phenotype. Furthermore, in analyses of 26 normal subjects studied on 2 separate occasions 7 days apart reported previously (12), the slope of the line relating total integrated esophageal acidity to total integrated gastric acidity was not significantly different from zero (results not shown).

**Figure 3.**
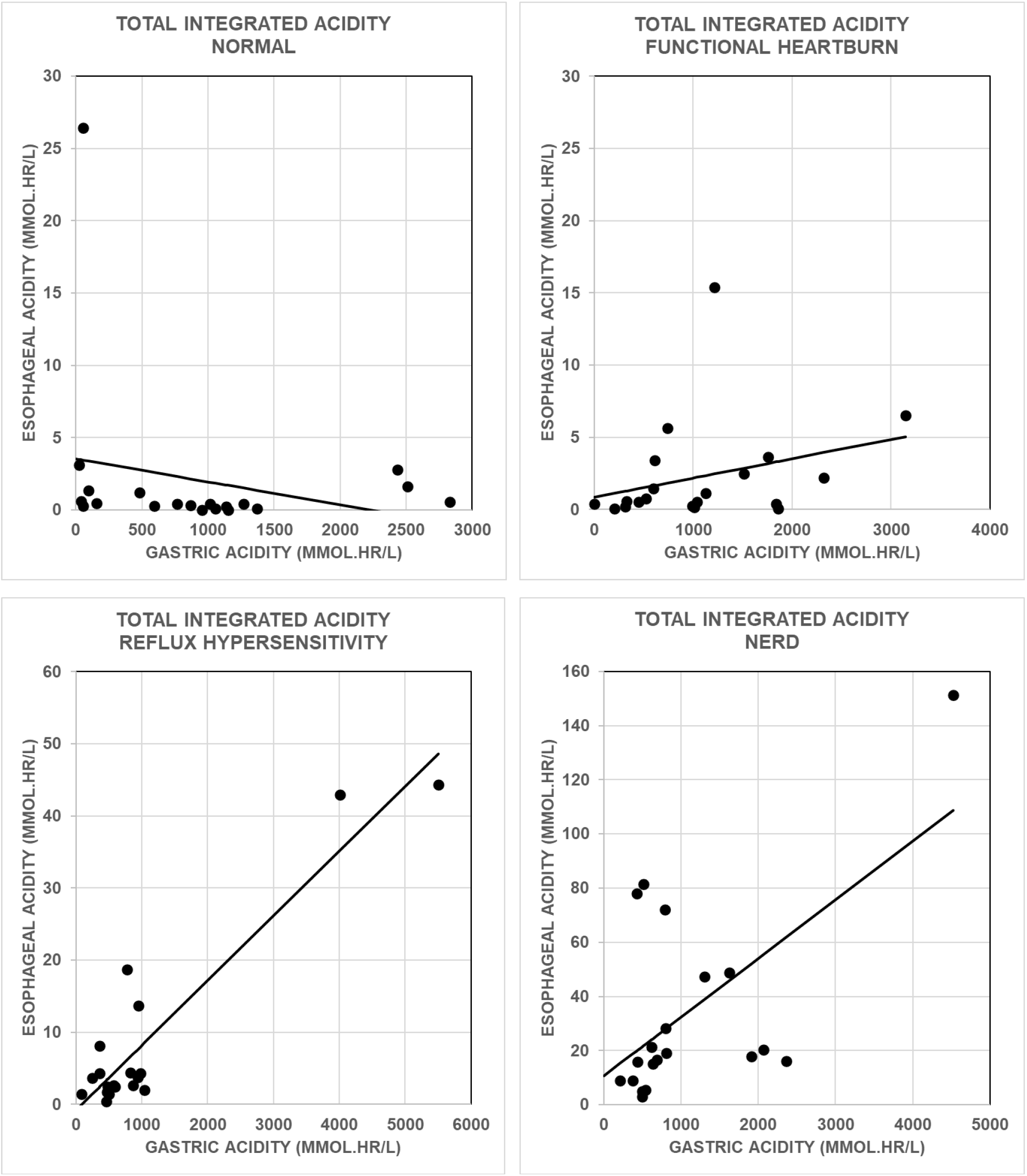
Plots of total integrated esophageal acidity versus total integrated gastric acidity for GERD phenotypes and normal subjects. Results are from 20 subjects in each GERD phenotype and from 19 normal subjects.

Results in Table 3 also show that the slope of the line relating total time esophageal pH<4 to total time gastric pH<4 in Figure 4 was not significantly different from zero and was not significantly non-linear for any GERD phenotype or Normal subjects De bartoli and colleagues (13) treated subjects with GERD symptoms and a negative upper gastrointestinal endoscopy with a single daily standard dose of a proton pump inhibitor (PPI) for 8 weeks. Subjects’ symptoms were assessed before and at the end of PPI treatment using the GERD impact scale (GIS) and a visual analog scale (VAS) for heartburn and regurgitation. A satisfactory response to the PPI was a decrease in the GIS and VAS of at least 75% from baseline. An unsatisfactory response was a decrease in the GIS and VAS of less than 50% from baseline. After stopping PPI treatment for 14 days, all subjects underwent impedance-pH measurements that were used to assign subjects to Functional Heartburn, Reflux Hypersensitivity or NERD groups.

**Table 3.** RESULTS FROM LINEAR REGRESSION ANALYSES OF ESOPHAGEAL ACIDITY VERSUS GASTRIC ACIDITY.

|  |  |  |  |  |
| --- | --- | --- | --- | --- |
|  | **NORMAL** | **FUNCTIONAL**<br>**HEARTBURN** | **REFLUX**<br>**HYPERSENSITIVITY** | **NERD** |
| <u>INTEGRATED ACIDITY</u> |  |  |  |  |
| SLOPE | -0.00158 | 0.00134 | 0.00898 | 0.0217 |
| R<sup>2</sup> | 0.0529 | 0.0870 | 0.8607 | 0.3583 |
| P-VALUE SLOPE<br>NON-ZERO | 0.3291 | 0.2069 | <0.0001 | 0.0053 |
| P-VALUE SLOPE<br>NON-LINEAR | 0.0227 | 0.460 | 0.208 | 0.570 |
| <u>TIME PH<4</u> |  |  |  |  |
| SLOPE | 0.0654 | 0.0143 | 0.0554 | 0.0788 |
| R<sup>2</sup> | 0.0846 | 0.106 | 0.173 | 0.0053 |
| P-VALUE SLOPE<br>NON-ZERO | 0.2134 | 0.1614 | 0.0685 | 0.7604 |
| P-VALUE SLOPE<br>NON-LINEAR | 0.8174 | 0.95551 | 0.8842 | 0.7604 |
| P-VALUE SLOPE NON-ZERO was determined with an F-test. P-VALUE SLOPE NON-LINEAR was determined with a Runs test. Results are from linear, least-squares regression analyses of the data in Figures 3 and 4. |  |  |  |  |

**Figure 4.**
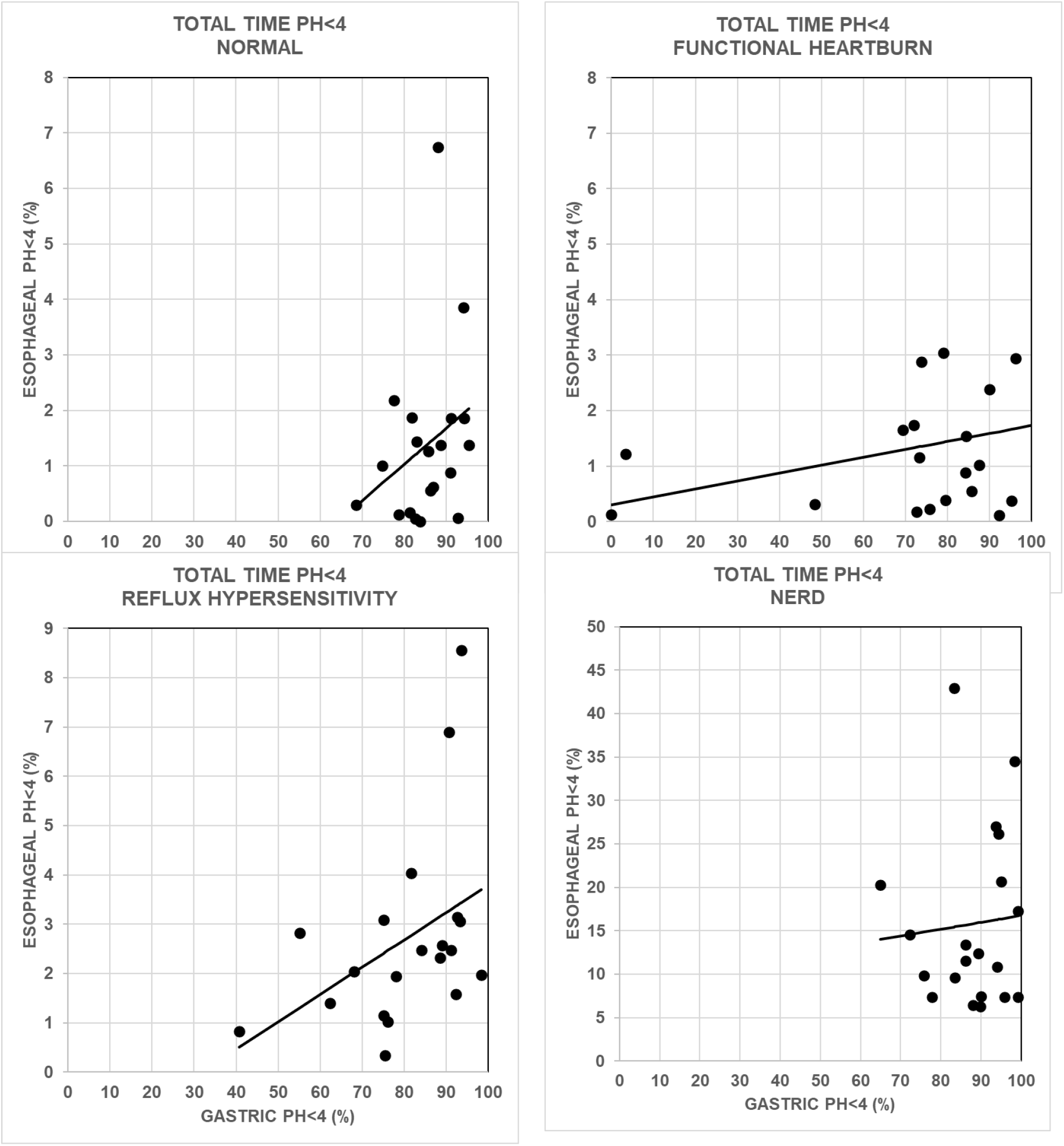
Plots of total time esophageal pH<4 versus total time gastric pH<4 for GERD phenotypes and normal subjects. Results are from 20 subjects in each GERD phenotype and from 19 normal subjects.

The magnitude of the slopes of the lines relating total integrated esophageal acidity to total integrated gastric acidity illustrated in Figure 3 measure the strength of the association between esophageal acidity and gastric acidity. Figure 5 compares the response as well as the non-response of a particular GERD phenotype to a PPI reported in reference 13 to the value of the slope for the same GERD phenotype given in Table 3. I included the slope for Functional Heartburn subjects in Figure 3 even though it was not significantly different from zero because it seemed possible that the lack of statistical significance for this slope might be due to an under-powered sample size.

**Figure 5.**
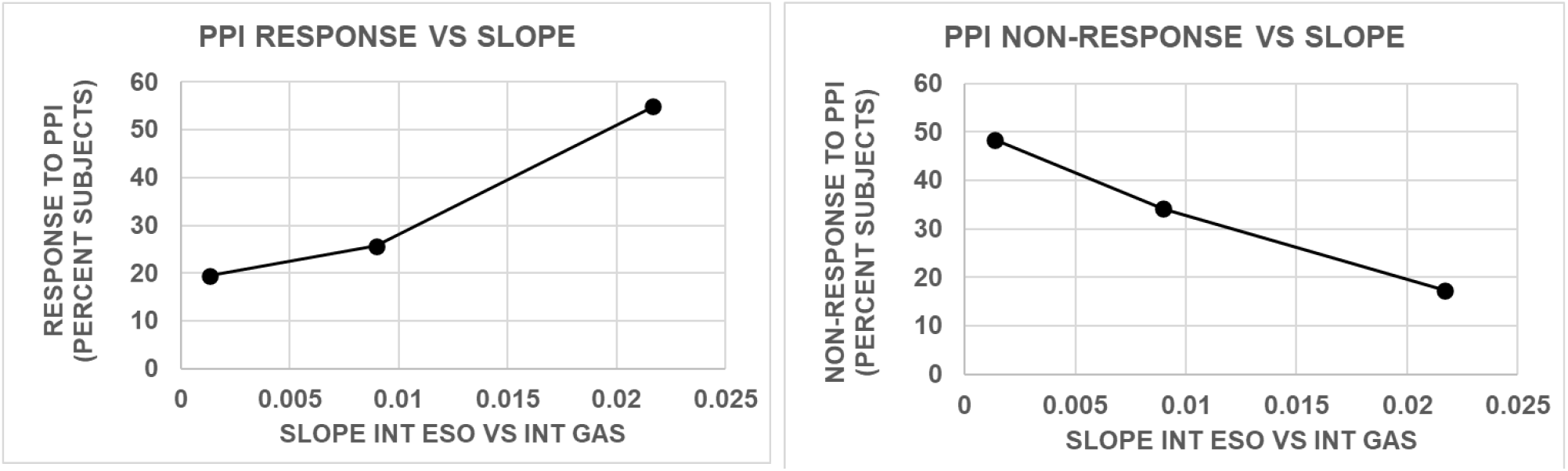
Plots of the percent of subjects responding to a proton pump inhibitor (PPI) and percent of subjects not responding to a PPI for an individual GERD phenotype and the corresponding slope of a plot of total integrated esophageal acidity (INT ESO) versus total integrated gastric acidity (INT GAS) for the same phenotype. Values for PPI responders and non-responders were calculated from data in Figure 3 in reference 13. Values for slopes were from data in Figure 3 and Table 3 from 20 subjects in each GERD phenotype.

Figure 5 shows that as the slope that quantifies the relationship between esophageal and gastric acidity increases, the proportion of subjects that respond to a PPI increases and the proportion of subjects that do not respond to a PPI decreases.

## DISCUSSION

Previous analyses of esophageal pH data from the same subjects that provided data for the present analyses found that the esophageal pH values followed a different power law distribution in each group of normal subjects and different GERD phenotypes. The distributions of changes in esophageal acid concentrations were greatest in normal subjects, less in Functional Heartburn subjects, still less in Reflux Hypersensitivity subjects and least in NERD subjects (1). The present analyses, however, found different distributions for gastric pH values that could be described by a 5^th^-order polynomial in normal subjects and each GERD phenotype, but virtually identical distributions of changes in gastric acid concentrations in the different groups of subjects. The difference between the distribution of gastric pH values and the distribution of changes in gastric acidity is at least partly attributable to distributions of changes in gastric acid concentration being adjusted for the values of acid concentration per se by calculating change with each sequential concentration expressed as a percentage of total cumulative gastric acid concentration. Figures 1 and 3 show the wide range of differences in gastric pH and acidity among the different subject groups that could influence the distributions of changes in acid concentration if not adjusted for. Others (10) have calculated the change in values in a time-series as a percentage difference from the mean of all values.

Since gastric acid is generally considered to be the source of esophageal acid in normal as well as GERD subjects, one might not expect differences between the distributions of esophageal pH and gastric pH or between the distributions of changes in esophageal acidity and gastric acidity. On the other hand, analyses using vector autoregression, a theory-free set of inter-related linear regressions used to measure relationships that can change over time, found that in pH records from normal subjects, as well as GERD subjects alone and after treatment with a proton pump inhibitor, gastric pH values provided important information regarding subsequent values of esophageal pH and values of esophageal pH provided important information regarding subsequent values of gastric pH (14). The ability of gastric pH and esophageal pH to provide information regarding subsequent values of each other was reduced in subjects with GERD compared to normal subjects. Furthermore, in normal subjects infusing acid into the esophagus, but not into the stomach, reduced gastric acidity (15). Infusing acid into the esophagus of normal subjects, however, did not alter gastric acidity during fasting and although the magnitude of the decrease in gastric acidity varied directly with meal-stimulated gastric acid secretion, the decrease in gastric acidity could only be detected beginning 3 hours after the end of the meal (15). These findings are consistent with the hypothesis that esophageal acidity and gastric acidity can influence each other in normal subjects and GERD phenotypes, and by so doing possibly account for the variation in the differences in the distributions of values of pH and acid concentrations among normal subjects and different GERD phenotypes.

There was a significant positive relationship between total integrated esophageal acidity and total integrated gastric acidity in NERD and Reflux Hypersensitivity subjects and possibly in Functional Heartburn subjects as well, but not in normal subjects. In contrast, there was no significant relationship between esophageal acidity and gastric acidity when acidity was measured as time pH<4. A previous study also found a poor correlation between integrated esophageal acidity and time esophageal pH<4 (12). The lack of a relationship between esophageal acidity and gastric acidity measured as time pH<4 is likely due to the fact that time gastric pH<4 truncates values of high gastric acidity. For example, one hour of gastric pH of 3.5 and one hour of gastric pH of 1.5 would have the same value of time pH<4 even though there is a 100-fold difference in the acid concentrations.

The strength of the relationship between esophageal acidity and gastric acidity reflected by the magnitude of the slope of a linear regression analysis of the two values in different GERD phenotypes correlated directly with the responses of different GERD phenotypes to PPI treatment and inversely with the lack of response to PPI treatment in the same phenotypes. It seems possible that the reduced variation in esophageal acid concentrations in a particular GERD phenotype results in tighter coupling with gastric acidity and the resulting effectiveness of PPI treatment that reduces esophageal acidity by acting on gastric acidity. One limitation of the present analyses, however, is that we have no data for the clinical effectiveness of PPIs from the subjects that provided the pH recordings and instead, relied on published results from other subjects.

Another limitation of the present analyses is that they do not provide an explanation for the existence of symptoms such as heartburn and regurgitation in Functional Heartburn and Reflux Hypersensitivity subjects but not in normal subjects although all three groups have normal esophageal acid exposure. Previous analyses of the same subjects used for the present study found that esophageal acid sensitivity appears to oscillate in each GERD phenotype, and for a given value of esophageal acid sensitivity, Reflux Hypersensitivity subjects have significantly more sequential symptoms associated with this sensitivity than do Functional Heartburn subjects (3). This difference between Functional Heartburn subjects and Reflux Hypersensitivity subjects might indicate that the lower variation of values of esophageal acid concentrations in Reflux Hypersensitivity subjects can account for the increase in GERD symptoms. The lower variation of esophageal acid concentrations in GERD phenotypes than in normal subjects might also explain why GERD phenotypes but not normal subjects have heartburn and regurgitation.

On the other hand, the oscillation of esophageal acid sensitivity in GERD phenotypes might reflect a property of esophageal mucosa that results in symptoms being produced by normal or increased esophageal acid exposure regardless of the variation in esophageal acid concentrations.

## Supporting information

COVER LETTER

## Data Availability

All data produced in the present study are available upon reasonable request to the authors.

## REFERENCES

1. Gardner JD. Decreased variation in esophageal acid concentration in different phenotypes of symptomatic gastroesophageal reflux disease. https://www.medrxiv.org/content/10.1101/2023.03.31.23288021.

2. Gardner JD. The relationship between esophageal acidity and symptom frequency in symptomatic nonerosive gastroesophageal reflux disease. Physiological Reports. 2022; 10, e15442.

3. Gardner JD. Oscillating esophageal acid sensitivity in symptomatic reflux hypersensitivity and functional heartburn. Qeios. 2022 doi:10.32388/IJUE1J.2.

4. Sifrim D, Roman S, Savarino E, Bor S, Bredenoord AJ, Castell D, Cicala M, De Bortoli N, Frazzoni M, Gonlachanvit S, Iwakiri K, Kawamura O, Krarup A, Lee YY, Ngiu CS, Ndebia E, Patcharatraku T, Pauwels A, De la Serna JP, Ramos R, Remes-Troche JM, Ribolsi M, Sammon A, Simren M,Tack J, Tutuian R, Valdovinos M Xiao, Zerbib F, Gyawali CP. Normal values and regional differences in oesophageal impedance-pH metrics: a consensus analysis of impedance-pH studies from around the world. Gut 2021; 70:1441–1449.

5. Gyawali CP, Kahrilas PJ, Savarino E, Zerbib F, Mion F, Smout Ajpm, Vaezi M, Sifrim D, Fox MR, Vela MF, Tutuian R, Tack J, Bredenoord AJ, Pandofilno J, Roman S. Modern diagnosis of GERD: the Lyon Consensus. Gut 2018; 67:1351–1362.

6. Ghisa M, Barberio B, Savarino V, Marabotto E, Ribolsi M, Bodini G, Zingone F, Frazzoni M, Savarino E. The Lyon Consensus: Does It Differ From the Previous Ones? J Neurogastroenterol Motil 2020;26:311–321.

7. Exceptions to Institutional Review Board requirements for research. United States Code of Federal Regulations, Title 45, Part 46, Subpart A, Criterion 4.

8. Weiner GJ, Richter JE, Copper JB, Wu WC, Castell DO. The symptom index: A clinically important parameter of 24-hour esophageal pH monitoring. Am J Gastroenterol 1988; 83:358–361.

9. Weusten BL, Roelofs JM, Akkermans LM, Van Berge-Henegouwen Gp, Smout AJ. The symptom-association probability: An improved method for symptom analysis of 24-hour esophageal pH data. Gastroenterology 1994; 107:1741–1745.

10. Fossion R, River AL, Estanol B. A physicist’s view of homeostasis: how time series of continuous monitoring reflect the function of physiological variables in regulatory mechanisms. Physiol Meas. 2018, 39, 084007.

11. Gardner JD, Perdomo C, Sloan S, Hahne WF, Barth JA, Rodriguez-Stanley S, Robinson M. Integrated acidity and rabeprazole pharmacology. Aliment Pharmacol Ther 2002; 16:455–464.

12. Gerson LB Triadafilopoulos G, Sahbaie P, Young W, Sloan S, Robinson M, Miner PB Jr., Gardner JD. Time esophageal pH < 4 overestimates the prevalence of pathologic esophageal reflux in subjects with gastroesophageal reflux disease treated with proton pump inhibitors. BMC Gastroenterology 2008, 8:15.

13. De bortoli, N, Martinucci I, Savarino E, Bellini M, Bredenoord AJ, Franchi R, Bertani L, Furnari M, Savarino V, Blandizzi C, Marchi S. Proton pump inhibitor responders who are not confirmed as GERD patients with impedance and pH monitoring: who are they? Neurogastroenterol Motil (2014) 26, 28–35.

14. Lu L, Mu JC, Sloan S, Miner PB Jr, Gardner JD. Exploring the physiologic role of human gastroesophageal reflux by analyzing time-series data from 24-h gastric and esophageal pH recordings. Physiol Rep, 20141; 2, e12051.

15. K. Blondeau K, Sifrim D, Gardner JD. Continuous distal oesophageal acidification decreases postprandial gastric acidity in healthy human subjects. Aliment Pharmacol Ther 2009; 29:561–570.

