## Supplementary material for "GASTRIC ACIDITY IN DIFFERENT PHENOTYPES OF SYMPTOMATIC GASTROESOPHAGEAL REFLUX DISEASE": COVER LETTER

The accompanying paper is based on analyses of data from the same subjects and using the same methods that I used for a previous paper cited below. As a result, the text describing the Subjects and Methods are the same in the two papers even though the Introduction, Results and Conclusions are different.

Gardner JD. Decreased variation in esophageal acid concentration in different phenotypes of symptomatic gastroesophageal reflux disease. https://www.medrxiv.org/content/10.1101/2023.03.31.23288021
